# Patterns in Medication Use for Treatment of Depression in Autistic Spectrum Disorder

**DOI:** 10.1101/2022.10.21.22280994

**Authors:** Riley A. Argonis, Ernest V. Pedapati, Kelli C. Dominick, Katherine Harris, Martine Lamy, Cara Fosdick, Lauren Schmitt, Rebecca C. Shaffer, Elizabeth Smith, Meredith Will, Christopher J. McDougle, Craig A. Erickson

## Abstract

Depression impacts many individuals with autism spectrum disorder (ASD), carrying increased risk of functional impairment, hospitalization, and suicide. Prescribing medication to target depression in patients with ASD occurs despite limited available systematic data describing medication management of depression in this population. Drawing from a large clinical database describing the prescribing practices in patients with ASD, we identified 170 individuals with ASD (mean age 16.2±8.7 years old) who received medication targeting symptoms of depression. We report prescribing rates for specific drugs, drug treatment duration, and reasons for drug discontinuation when applicable. Sertraline, mirtazapine, and fluoxetine were the three most commonly prescribed medications to treat comorbid depression for this patient population. Among 247 drug starts, 121 (49%) drug treatments were continued at the final reviewed follow-up visit (average treatment duration of ±0.72 years). The most common reason for discontinuation across all medications prescribed was loss of or lack of effectiveness. This study raises concern that standard of care pharmacological treatments for depression in individuals with ASD may be less effective than in neurotypical populations. There remains a need to develop effective interventions for depression specifically tailored to the needs of individuals with ASD.

## Introduction

Mental illness is a growing concern among adolescents and adults alike, with 21% of adults and 49.5% of adolescents aged 13-17 years in the United States experiencing any mental disorder in the past year (NIMH, 2022). Autistic individuals may be at particular risk for mental health concerns, with over 2 in 3 autistic adults (Fombonne, et al., 2020) and up to 80% of autistic adolescents (Buck, et al., 2014) diagnosed with at least one comorbid psychiatric disorder (Fombonne, et al., 2020). Depression disproportionately affects individuals with autism spectrum disorder (ASD) and contributes to functional disability, including educational and vocational impairment, social withdrawal, and suicide (Cassidy et al., 2014). The risk of developing major depressive disorder (MDD) is nearly four times greater in individuals with ASD than in their neurotypical peers (Hudson, et.al., 2018). Other studies have found rates of depression in autistic patients higher than rates in the general population, including rates of depression of 23% of autistic adults (Hollacks, et al., 2018) and 30% of autistic adolescents (Maton & Nebel-Schwalm, 2006) compared to rates of 8% and 17% of adults and adolescents respectively of depression (NIMH, 2022) in the general population. Understanding the true prevalence rates of depression in the autistic population is difficult due in part to a lack of validated instruments and measures to identify atypical presentations of comorbid depression in individuals with ASD (DeFilippis, 2018). The presence or absence of intellectual disability (ID) in the context of ASD may also impact rates of depression noted among autistic individuals. Individuals with ASD without an ID are more likely to be diagnosed with comorbid depression (Buck, et al., 2014; Hudson et al., 2019). This may be related to a tendency to exhibit canonical signs and symptoms of MDD, including sadness, anhedonia, cognitive changes, suicidality, and neurovegetative symptoms among the subgroup of persons with ASD and higher cognitive ability. Moreover, depressive symptoms can also present atypically in ASD and have overlapping features with ASD (e.g., social withdrawal and flat affect). These symptoms may include increased irritability (including self-injury), loss of special interests, engagement in repetitive behavior, and decreases in adaptive function and self-care (Pezzimenti et al., 2019). Higher rates of the diagnosis of MDD in autistic individuals without ID may also be due to their relative language ability, with these individuals more able to communicate thoughts and feelings, given that diagnosing depression often relies on patient self-report (Buck, et al., 2014). Therefore, MDD may be underdiagnosed in autistic individuals with ID and/or significant expressive language deficits (Cassidy et al., 2014; Cassidy & Rogers, 2017; Magnuson & Constantino, 2011).

Not surprisingly, given high rates of comorbid mental illness in autistic patients, reports have noted high psychiatric medication use in this population with a high rate of polypharmacy. In a large registry of autistic children, Rosenberg et al. (2009) noted that 35% of youth received at least one psychotropic drug targeting mental illness and 10% received at least three different classes of psychiatric drugs at one time (Rosenberg, et al., 2009). A second report noted 40% of autistic youth took at least one psychiatric drug targeting comorbid mental illness, and up to 31% received at least two psychiatric medications (Feroe, et al., 2021). Specific to depression in autistic adults, Zheng et al. 2021 noted among all patients receiving medicating, psychotherapy, or both, 85% received a psychotropic medication targeting symptoms of depression (Zheng, et al., 2021). Despite the frequency of psychiatric medication use targeting comorbid mental illness in autistic patients, little information is available describing specific drug use, dosing, and tolerability to inform this prescribing practice. This is particularly true regarding the medication management of depression in autistic patients.

In this study, we aim to extract information on specific prescribing patterns targeting symptoms of depression in autistic patients from a large clinical medication management database focused on this population. We will review specific medications utilized, medication dosing, duration of treatment, and aspects of drug tolerability, including whether specific drug trials were maintained long-term.

## Methods

### Procedure

Data for this report was gathered from a well-established patient research database (Wink et al., 2017; Dominick et al., 2015; Adler et al., 2015; Wink et al., 2014) describing clinical medication management in 1100 autistic patients treated at an academic medical center from July 2004 to April 2012. The database was queried for autistic patients who received one or more medications targeting symptoms of depression associated with an established diagnosis of MDD.

Descriptive statistics of demographic information and medication usage patterns were generated using the statistical software JASP (version 0.16; Amsterdam, Netherlands) to summarize the sample. Demographic variables include age, sex assigned at birth, and race. Additional variables included the presence or absence of ID, the presence or absence of Fragile X syndrome, and the specific ASD-associated diagnosis using DSM-IV-TR classification: namely, Autistic Disorder, Pervasive Developmental Disorder Not Otherwise Specified (PDD-NOS), or Asperger’s Disorder.

For each medication associated with targeting depression, we captured the generic medication name, duration of treatment across visits, and reported side effects. We assigned each medication to one of five drug class categories including selective serotonin reuptake inhibitor (SSRI), serotonin-norepinephrine reuptake inhibitor (SNRI), antipsychotic (AP), other serotonin modulator (OSM), and other class (OC). Patients who received more than one medication for depression at the same time were also noted. If prescription of a drug was recorded through the patient’s final visit in the database, this was counted as a continued medication. If a medication was discontinued, the reason for discontinuation (if available) was noted. The mean and standard deviation for drug dosing at the last recorded visit was calculated. Infrequently prescribed medications (less than five times) are summarized in Supplementary Tables.

## Results

### Patient Demographics

One hundred and seventy patients (15.5% of the total database sample) had received at least one medication targeting symptoms of depression in the context of a diagnosis of MDD or depressive disorder NOS. Our sample of 170 autistic patients had a mean age of 16.2 years (SD= 8.7 years) with an age range of 2.6-78.8 years. Consistent with prevalence estimates of ASD, males outnumbered females in our sample at a ratio of 4.8 to 1. The sample was overwhelmingly white, with 4.3% of the patient sample being black. Eighteen (10.5%) of the patients had Fragile X syndrome-associated autism. Among specific autism-associated diagnoses made at the time using DSM-IV-TR, 72 (41.9%) were diagnosed with autistic disorder, 66 (38.4%) with PDD-NOS; and 32 (18.6%) with Asperger’s disorder. Eighty-one (47.6%) of the patients had a comorbid diagnosis of ID. See Table 1 for summary of demographic and clinical details.

**Table 1:**
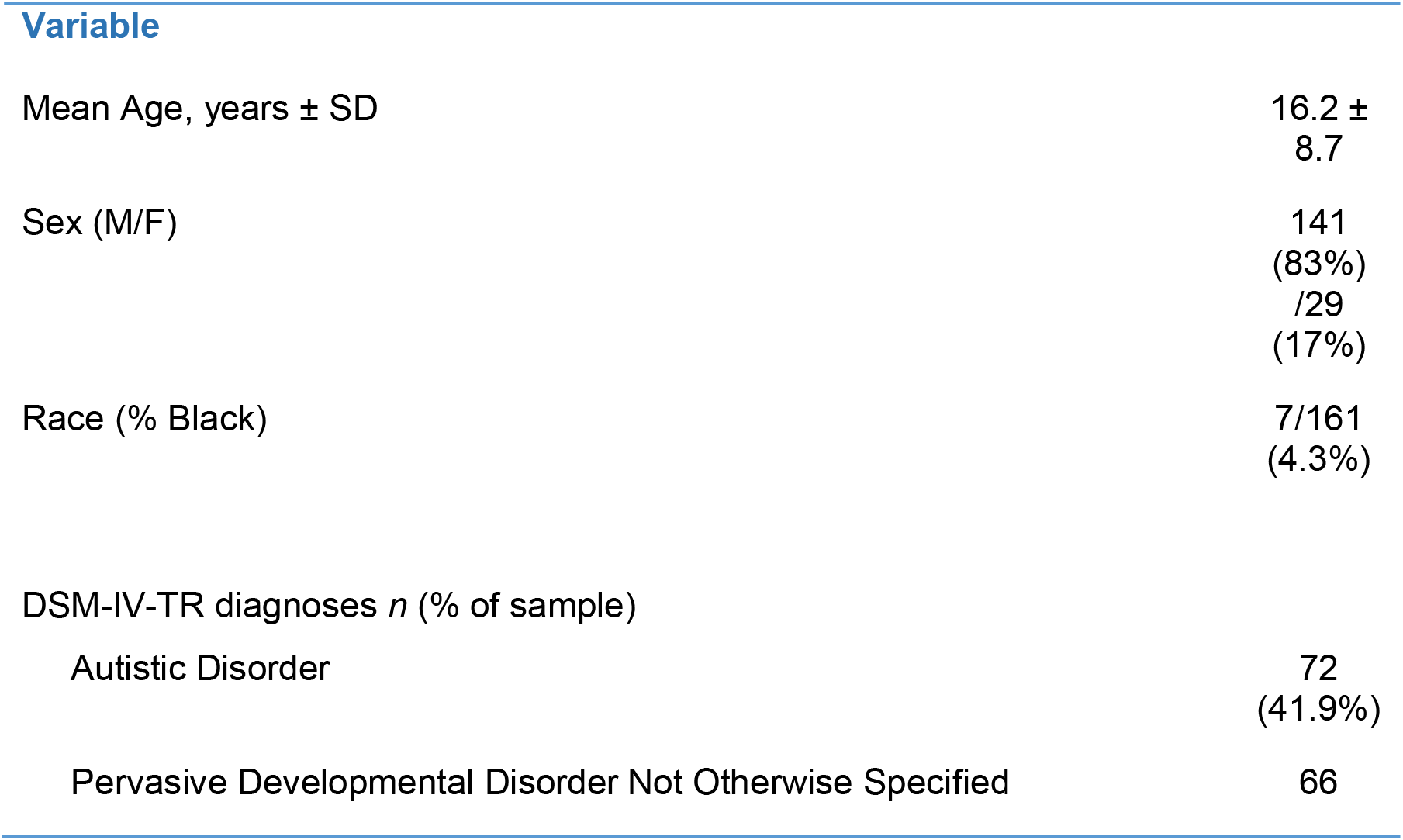
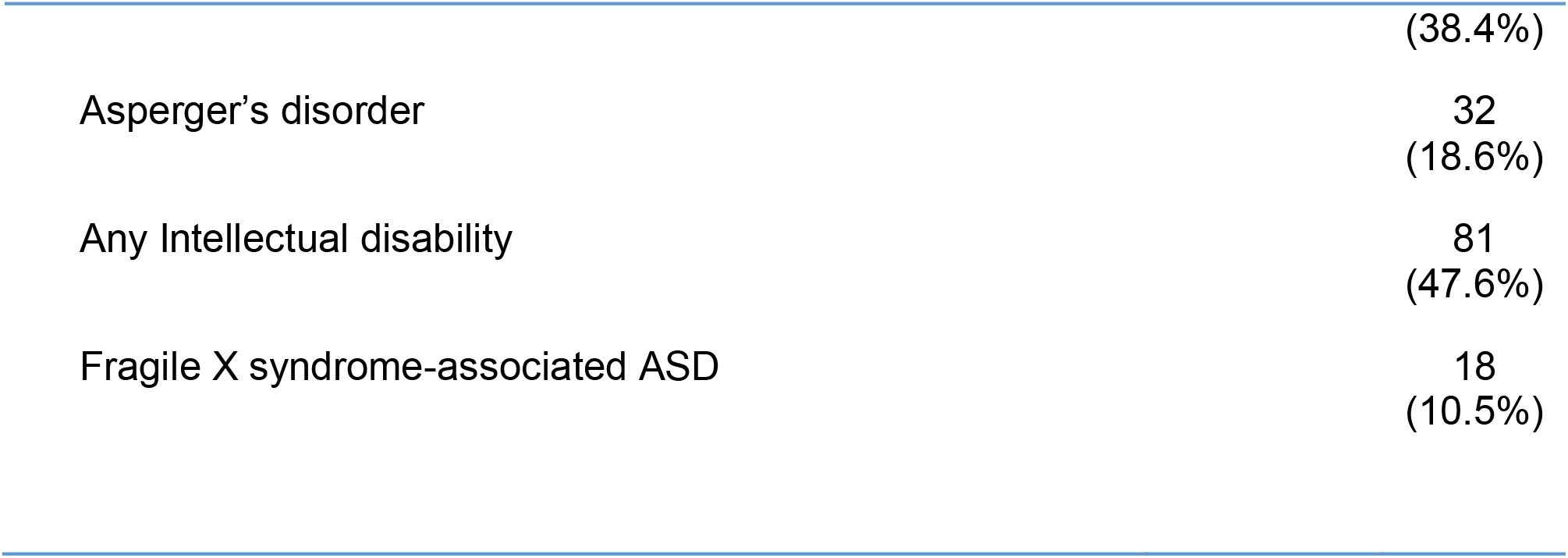
Characteristics of Study Sample (*N*=170)

| Variable |  |
| --- | --- |
| Mean Age, years ± SD | 16.2 ± 8.7 |
| Sex (M/F) | 141 (83%)<br>/29 (17%) |
| Race (% Black) | 7/161 (4.3%) |
| DSM-IV-TR diagnoses *n* (% of sample) |  |
| Autistic Disorder | 72 (41.9%) |
| Pervasive Developmental Disorder Not Otherwise Specified | 66 |
|  | (38.4%) |
| Asperger's disorder | 32<br>(18.6%) |
| Any Intellectual disability | 81<br>(47.6%) |
| Fragile X syndrome-associated ASD | 18<br>(10.5%) |

### Medication Use Details

In total, 23 different medications were utilized to treat depression in our patient cohort. The use of two drugs (L-methylfolate and fluvoxamine) did not have valid data for dosage or duration of use, and therefore these medications were excluded from our final data analysis. The remaining 21 medications accounted for over 247 medication starts within the cohort. Of these, 11 medications were prescribed more than five times (see Table 2) and served as the main focus of this analysis. Among these 11 medications, five are SSRIs, two are SNRIs, one is an antipsychotic, two are OSMs, and one is from another class. Medications that were prescribed five or fewer times are summarized in supplementary tables. Twenty-six patients (15.3%) received two or more medications at one time to treat depression.

**Table 2:** Details of Medication Use

| Medication | Mean final daily dose, ± SD (mg) | Mean age of drug initiation (years) | Mean duration of treatment (years) |
| --- | --- | --- | --- |
| Sertraline (n=70) | 91.2 ± 58.3 | 15.1 | 1.2 |
| Mirtazapine (n=32) | 21.8 ± 13.8 | 13.7 | 0.9 |
| Fluoxetine (n=30) | 26.7 ± 20.6 | 13.5 | 1.0 |
| Bupropion (n=21) | 241.8 ± 95.2 | 18.3 | 0.6 |
| Citalopram (n=19) | 31.1 ± 20.0 | 15.0 | 1.1 |
| Escitalopram (n=14) | 15.1 ± 7.8 | 17.8 | 0.9 |
| Duloxetine (n=14) | 61.3 ± 35.7 | 22.1 | 0.5 |
| Paroxetine (n=13) | 23.5 ± 15.1 | 15.6 | 0.7 |
| Aripiprazole (n=7) | 10.4 ± 6.7 | 16.2 | 0.7 |
| Venlafaxine (n=6) | 112.5 ± 99.7 | 23.7 | 0.7 |
| Trazodone (n=6) | 150 ± 122.8 | 15.8 | 0.2 |

The three most prescribed drugs were sertraline, mirtazapine, and fluoxetine. Sertraline was the most prescribed drug to patients (n=70 patients receiving sertraline at some point) representing 20% of all medication uses in this sample. Patients were, on average, 15.1 years old when first prescribed sertraline, and the average final dose was 91.2 mg per day (*SD* = 58.3 mg). Thirty two patients continued on sertraline at the last visit (45.7% continuation rate). Mirtazapine was the second-most prescribed medication to treat depression with 32 prescription starts (nearly 13% of all medication uses). Patients were, on average, 13.7 years old at the beginning of their treatment with mirtazapine, and the average final dose prescribed was 21.8 mg per day (*SD*=13.8 mg). Eleven patients continued on mirtazapine at the final available visit (34.4% continuation rate). Fluoxetine was prescribed 30 times, the third most commonly prescribed drug (12% of all medication uses). The average age at start of fluoxetine treatment was 13.5 years old, and the average final recorded daily dose was 26.7 mg (*SD*=20.6 mg). Sixteen patients continued on fluoxetine at the last visit (53.3% continuation rate).

### Medication Discontinuation Details

Reasons for discontinuation of the 11 most commonly prescribed medications are tabulated in Table 3. Overall, “loss of effectiveness” was by far the most common reason for medication discontinuation. Other reasons for the discontinuation of a drug across the 11 drugs of interest were “aggression,” “weight gain,” and “sedation/tiredness.” At times, no reason was given in the data for discontinuation. Among all 247 medication trials reviewed in this analysis, 121 drug treatments were continued at the final visit (49% overall drug continuation rate). For those drug trials discontinued, the average duration prior to discontinuation was 0.72 years.

**Table 3:** Details of Drug Discontinuation

| Medication | Total medication starts | Continuations at last recorded follow-up visit (% of starts) | Most common reasons for drug discontinuation | n = X, (% of starts) |
| --- | --- | --- | --- | --- |
| Sertraline | 70 | 32 (45.7%) | Loss of effectiveness | 6 (8.6%) |
|  |  |  | Aggression | 3 (4.3%) |
|  |  |  | Study | 2 (2.9%) |
| Mirtazapine | 32 | 11 (34.4%) | Loss of effectiveness | 5 (15.6%) |
|  |  |  | Weight gain | 4 (12.5%) |
|  |  |  | Sedation/tiredness | 2 (6.3%) |
| Fluoxetine | 30 | 16 (53.3%) | Loss of effectiveness | 8 (26.7%) |
|  |  |  | Sedation/tiredness | 1 (3.3%) |
|  |  |  | Tics | 1 (3.3%) |
| Bupropion | 21 | 14 (66.7%) | Loss of effectiveness | 1 (4.8%) |
|  |  |  | Suicidality | 1 (4.8%) |
|  |  |  | Weight gain | 1 (4.8%) |
| Citalopram | 19 | 10 (52.6%) | Loss of effectiveness | 3 (15.8%) |
|  |  |  | Sedation/tiredness | 2 (10.5%) |
|  |  |  | Confusion/disorientation | 1 (5.3%) |
| Escitalopram | 14 | 6 (42.9%) | Loss of effectiveness | 5 (35.7%) |
| Duloxetine | 14 | 7 (50%) | Loss of effectiveness | 6 (42.9%) |
|  |  |  | Increased repetitive behavior | 1 (7.1%) |
| Paroxetine | 13 | 8 (61.5%) | Loss of effectiveness | 1 (7.7%) |
|  |  |  | Study | 1 (7.7%) |
|  |  |  | Weight gain | 1 (7.7%) |
| Aripiprazole | 7 | 4 (57.1%) | Loss of effectiveness | 1 (14.3%) |
|  |  |  | Sedation/tiredness | 1 (14.3%) |
| Venlafaxine | 6 | 3 (50%) | Discontinued by primary<br>care physician | 1 (16.7%) |
| Trazodone | 6 | 5 (83.3%) | Difficulty swallowing | 1 (16.7%) |

## Discussion

Here we summarize patterns of medication use for depression in a large clinical cohort of individuals with ASD. Given that no randomized controlled medication trials focused on treating depression in ASD have been published, we expected prescribing patterns to vary and likely be based on evidence-based practices in non-ASD populations (Kim & Lecavalier, 2021; McCracken et al., 2021; Pezzimenti et al., 2019). In our sample, we found that medications targeting depression were discontinued around half the time, usually based on perceived lack of effectiveness. This is consistent with the growing observations that treatment response to standard-of-care medications for mood disorders may be unpredictable or they may be poorly tolerated in individuals with ASD (Masi et al., 2017).

Our data, like others, found a high rate of prescribing SSRIs (Esbensen, et al., 2009). This is not unexpected, as SSRIs are commonly used to treat MDD in neurotypical populations. However, in ASD, SSRIs have been extensively studied outside of depression for core and non-core symptoms such as self-injury. A Cochrane review did not find any evidence of benefit (and potential emerging evidence of harm) for SSRI use in ASD (Williams et al., 2013). Our dataset precludes any objective analysis of clinical outcomes and does not support any clear consensus on effectiveness of any particular medication. Selective serotonin reuptake inhibitors were used for the longest duration of time in our study, with sertraline, fluoxetine, and citalopram being the only medications to be taken for a year or longer. On average, SSRI use in our dataset was initiated during adolescence. This pattern of use differed from that of SNRIs in our dataset, where venlafaxine, desvenlafaxine, and duloxetine were most often initially prescribed to individuals in young adulthood (range of mean ages for SNRI initiation= 19.0-23.7 years).

Despite the pressing need, limited results from intervention research specifically targeting depression and suicidality in ASD is available. Thus, there is great interest in developing effective interventions for MDD specifically tailored to the needs of individuals with ASD.

In our analysis, one in two drug starts to treat depression resulted in drug discontinuation, which was most often due to lack of drug effectiveness. The reasons for this are likely multifactorial and will require alternative study designs to probe further. For example, lack of effectiveness may be associated with disease severity including severity of underlying ASD or depressive symptoms. Given higher prevalence rates for depression, suicidality, and polypharmacy in ASD, the rate of treatment-resistant MDD (following the failure of standard-of-care) in ASD may exceed that of the general population (Zhdanava et al., 2022). Our results may also ultimately underreport the rate of drug discontinuation, given the finite time frame and assumption that medications were continued if found on the final recorded visit in our dataset.

There are several limitations to our report in addition to those commonly associated with retrospective studies. First, we did not attempt to focus on treatment indications beyond depression within our cohort. For example, participants may have suffered from co-occurring anxiety disorders (reported to occur in 40% of autistic youth; White et. al., 2009), which would confound the interpretation of medication use. We may infer, for example, that the high frequency use of the antidepressant mirtazapine may reflect prescribing patterns to target specific use for anxiety in ASD (McDougle et al., 2022). It is also possible that mirtazapine use could have been preferred in subjects with depressive symptoms and associated insomnia given the potential benefits of this drug for sleep disturbance.

As mentioned earlier, the dataset does not link drug discontinuation with clinical outcomes. Thus, we are unable to assess if medication use led to a remission of symptoms of depression. Though lack of effectiveness was clearly stated as a reason for discontinuation, our data set did not allow us to determine if symptom remission was a reason for treatment discontinuation. While the dataset was collected at a research-oriented tertiary care autism center, the diagnoses of ASD represent clinical opinions of the prescribers based upon DSM-IV-TR criteria. It is unknown if diagnoses were externally validated with diagnostic instruments such as the Autism Diagnostic Observation Schedule (ADOS).

There is a clear need for more rigorous studies of depression in this patient population. While this may include prospective drug trials with rigorous patient characterization and use of validated outcome measures, it is also important to evaluate non-drug therapies for this indication including various psychotherapies and neuromodulation, among other modalities. Given our report, it appears that half of the time a medication is started targeting depression in autistic patients, the drug will be discontinued, most often due to lack of effectiveness. Due to the substantial burden depression places on the individual, family, and community, the need to establish more effective and well-studied treatments for depression in this population are required.

## Data Availability

All data produced in the present study are available upon reasonable request to the authors.

---

**Supplemental Table 1:** Details of Medication Use

| Medication | Mean final daily dose ± SD (mg) | Mean age of drug indication (years) | Mean duration (years if n>1) |
| --- | --- | --- | --- |
| Ziprasidone (n=3) | 33.3 ± 11.5 | 18.7 | 0.1 |
| Quetiapine (n=2) | 250 ± 70.7 | 15.9 | 0.3 |
| Imipramine (n=2) | 75 ± 35.4 | 13.8 | 1.7 |
| Desvenlafaxine (n=2) | 50 ± 0 | 19.0 | 0.1 |
| Olanzapine (n=1) | 20 | 26.1 | - |
| Methylphenidate (n=1) | 90 | 12.8 | - |
| Amphetamine and Dextroamphetamine. (n=1) | 5 | 10.2 | 0.2 |
| Clomipramine (n=1) | 125 | 12.5 | - |
| Lamotrigine (n=1) | 200 | 19.8 | 0.1 |
| Melatonin (n=1) | 6 | 12.2 | 0.3 |

**Supplemental Table 2:** Details of Drug Discontinuation

| Medication | Total medication starts | Continuations at last recorded follow-up visit (% of starts) | Most common reasons for drug discontinuation (n>1 discontinuation) |
| --- | --- | --- | --- |
| Ziprasidone | 3 | 0 (0%) | Increased anxiety |
| Quetiapine | 2 | 0 (0%) | Loss of effectiveness<br>Reduced expressive language |
| Imipramine | 2 | 1 (50%) | - |
| Desvenlafaxine | 2 | 1 (50%) | - |
| Olanzapine | 1 | 1 (100%) | - |
| Methylphenidate | 1 | 1 (100%) | - |
| Amph. and Dextroamph. | 1 | 0 (0%) | - |
| Clomipramine | 1 | 1 (100%) | - |
| Lamotrigine | 1 | 0 (0%) | - |
| Melatonin | 1 | 0 (0%) | - |

## References

Adler, B. A., Wink, L. K., Early, M., Shaffer, R., Minshawi, N., McDougle, C. J., & Erickson, C. A. (2015). Drug-refractory aggression, self-injurious behavior, and severe tantrums in autism spectrum disorders: A chart review study. Autism, 19(1), 102–106. https://doi.org/10.1177/1362361314524641

Bolton, P. F., Pickles, A., Murphy, M., & Rutter, M. (1998). Autism, affective and other psychiatric disorders: Patterns of familial aggregation. Psychological Medicine, 28(2), 385–395. https://doi.org/10.1017/s0033291797006004

Buck, T. R., Viskochil, J., Farley, M., Coon, H., McMahon, W. M., Morgan, J., & Bilder, D. A. (2014). Psychiatric comorbidity and medication use in adults with autism spectrum disorder. Journal of Autism and Developmental Disorders, 44(12), 3063–3071. https://doi.org/10.1007/s10803-014-2170-2

Cassidy, S., & Rodgers, J. (2017). Understanding and prevention of suicide in autism. The Lancet Psychiatry, 4(6). https://doi.org/10.1016/s2215-0366(17)30162-1

Cassidy, S., Bradley, P., Robinson, J., Allison, C., McHugh, M., & Baron-Cohen, S. (2014). Suicidal ideation and suicide plans or attempts in adults with Asperger’s syndrome attending a Specialist Diagnostic Clinic: A clinical cohort study. The Lancet Psychiatry, 1(2), 142–147. https://doi.org/10.1016/s2215-0366(14)70248-2

Cross-Disorder Group of the Psychiatric Genomics Consortium. (2013). Identification of risk loci with shared effects on five major psychiatric disorders: A genome-wide analysis. The Lancet, 381(9875), 1371–1379. https://doi.org/10.1016/s0140-6736(12)62129-1

DeFilippis, M. (2018). Depression in children and adolescents with autism spectrum disorder. Children, 5(9), 112. https://doi.org/10.3390/children5090112

Dominick, K., Wink, L. K., McDougle, C. J., & Erickson, C. A. (2015). A retrospective naturalistic study of ziprasidone for irritability in youth with autism spectrum disorder. Journal of Child and Adolescent Psychopharmacology, 25(5), 397–401. https://doi.org/10.1089/cap.2014.0111

Esbensen, A. J., Greenberg, J. S., Seltzer, M. M., & Aman, M. G. (2009). A longitudinal investigation of psychotropic and non-psychotropic medication use among adolescents and adults with autism spectrum disorders. Journal of Autism and Developmental Disorders, 39(9), 1339–1349. https://doi.org/10.1007/s10803-009-0750-3

Feroe, A. G., Uppal, N., Gutiérrez-Sacristán, A., Mousavi, S., Greenspun, P., Surati, R., Kohane, I. S., & Avillach, P. (2021). Medication use in the management of comorbidities among individuals with autism spectrum disorder from a large nationwide insurance database. JAMA Pediatrics, 175(9), 957–965. https://doi.org/10.1001/jamapediatrics.2021.1329

Fombonne, E., Green Snyder, L. A., Daniels, A., Feliciano, P., & Chung, W. (2020). Psychiatric and medical profiles of autistic adults in the spark cohort. Journal of Autism and Developmental Disorders, 50(10), 3679–3698. https://doi.org/10.1007/s10803-020-04414-6

Hollocks, M. J., Lerh, J. W., Magiati, I., Meiser-Stedman, R., & Brugha, T. S. (2018). Anxiety and depression in adults with autism spectrum disorder: A systematic review and meta-analysis. Psychological Medicine, 49(4), 559–572. https://doi.org/10.1017/s0033291718002283

Hudson, C. C., Hall, L., & Harkness, K. L. (2018). Prevalence of depressive disorders in individuals with autism spectrum disorder: A meta-analysis. Journal of Abnormal Child Psychology, 47(1), 165–175. https://doi.org/10.1007/s10802-018-0402-1

Kim, S. Y., & Lecavalier, L. (2021). Depression in young autistic people: A scoping review. Research in Autism Spectrum Disorders, 88, 101841. https://doi.org/10.1016/j.rasd.2021.101841

Magnuson, K. M., & Constantino, J. N. (2011). Characterization of depression in children with autism spectrum disorders. Journal of Developmental & Behavioral Pediatrics, 32(4), 332–340. https://doi.org/10.1097/dbp.0b013e318213f56c

Masi, A., DeMayo, M. M., Glozier, N., & Guastella, A. J. (2017). An overview of autism spectrum disorder, heterogeneity and treatment options. Neuroscience Bulletin, 33(2), 183–193. https://doi.org/10.1007/s12264-017-0100-y

Matson, J. L., & Nebel-Schwalm, M. S. (2007). Comorbid psychopathology with autism spectrum disorder in children: An overview. Research in Developmental Disabilities, 28(4), 341–352. https://doi.org/10.1016/j.ridd.2005.12.004

McCracken, J. T., Anagnostou, E., Arango, C., Dawson, G., McPartland, J., Murphy, D., Pandina, G., & Veenstra-VanderWeele, J. (2021). Corrigendum to “drug development for autism spectrum disorder (ASD): Progress, challenges, and future directions.” European Neuropsychopharmacology, 50, 133–134. https://doi.org/10.1016/j.euroneuro.2021.07.090

McDougle, C. J., Thom, R. P., Ravichandran, C. T., Palumbo, M. L., Politte, L. C., Mullett, J. E., Keary, C. J., Erickson, C. A., Stigler, K. A., Mathieu-Frasier, L., & Posey, D. J. (2022). A randomized double-blind, placebo-controlled pilot trial of Mirtazapine for anxiety in children and adolescents with autism spectrum disorder. Neuropsychopharmacology, 47(6), 1263–1270. https://doi.org/10.1038/s41386-022-01295-4

Pezzimenti, F., Han, G. T., Vasa, R. A., & Gotham, K. (2019). Depression in youth with autism spectrum disorder. Child and Adolescent Psychiatric Clinics of North America, 28(3), 397–409. https://doi.org/10.1016/j.chc.2019.02.009

Rosenberg, R. E., Mandell, D. S., Farmer, J. E., Law, J. K., Marvin, A. R., & Law, P. A. (2009). Psychotropic medication use among children with autism spectrum disorders enrolled in a National Registry, 2007–2008. Journal of Autism and Developmental Disorders, 40(3), 342–351. https://doi.org/10.1007/s10803-009-0878-1

Spencer, D., Marshall, J., Post, B., Kulakodlu, M., Newschaffer, C., Dennen, T., Azocar, F., & Jain, A. (2013). Psychotropic medication use and polypharmacy in children with autism spectrum disorders. Pediatrics, 132(5), 833–840. https://doi.org/10.1542/peds.2012-3774

U.S. Department of Health and Human Services. (2022, January). Major depression. National Institute of Mental Health. Retrieved August 1, 2022, from https://www.nimh.nih.gov/health/statistics/major-depression#part_2562

U.S. Department of Health and Human Services. (2022, January). Mental Illness. National Institute of Mental Health. Retrieved August 1, 2022, from https://www.nimh.nih.gov/health/statistics/mental-illness

White, S. W., Oswald, D., Ollendick, T., & Scahill, L. (2009). Anxiety in children and adolescents with autism spectrum disorders. Clinical Psychology Review, 29(3), 216–229. https://doi.org/10.1016/j.cpr.2009.01.003.

Wink, L. K., Early, M., Schaefer, T., Pottenger, A., Horn, P., McDougle, C. J., & Erickson, C. A. (2014). Body mass index change in autism spectrum disorders: Comparison of treatment with risperidone and Aripiprazole. Journal of Child and Adolescent Psychopharmacology, 24(2), 78–82. https://doi.org/10.1089/cap.2013.0099

Wink, L. K., Pedapati, E. V., Horn, P. S., McDougle, C. J., & Erickson, C. A. (2017). Multiple antipsychotic medication use in autism spectrum disorder. Journal of Child and Adolescent Psychopharmacology, 27(1), 91–94. https://doi.org/10.1089/cap.2015.0123

Zhdanava, M., Pilon, D., Ghelerter, I., Chow, W., Joshi, K., Lefebvre, P., & Sheehan, J. J. (2021). The prevalence and national burden of treatment-resistant depression and major depressive disorder in the United States. The Journal of Clinical Psychiatry, 82(2). https://doi.org/10.4088/jcp.20m13699

Zheng, S., Taylor, J. L., Adams, R., Pezzimenti, F., & Bishop, S. L. (2021). Perceived helpfulness of depression treatments among young adults with autism. Autism Research, 14(7), 1522–1528. https://doi.org/10.1002/aur.2515

